# Posttraumatic Stress Disorder and the Nature of Trauma in Patients with Cardiovascular Diseases: A Case-Control study

**DOI:** 10.1101/2021.01.14.21249762

**Authors:** Samuel Cyr, Marie-Joëlle Marcil, Valérie Long, Corrado De Marco, Katia Dyrda, Judith Brouillette

**Affiliations:** Research Centre, Montreal Heart Institute and Université de Montréal, Montreal, Quebec, Canada; Faculty of Pharmacy, Université de Montréal, Montreal, Quebec, Canada; Department of Psychiatry and Addiction, Faculty of Medicine, Université de Montréal, Montreal, Quebec, Canada; Department of Medicine, Faculty of Medicine, Université de Montréal, Montreal, Quebec, Canada

**Keywords:** Stress Disorders, Post-Traumatic, Cardiovascular Diseases, Medical Traumas

## Abstract

**Introduction:** A large body of evidence indicates a significant and morbid association between posttraumatic stress disorder (PTSD) and cardiovascular disease (CVD). Few studies, however, have addressed the range of trauma in this medical population, from massive heart attack, to defibrillator shock to previous interpersonal aggression.

**Objective:** The main objective of this study was to examine the nature of trauma associated with the development of PTSD in CVD patients. More precisely, we were interested in knowing if trauma was medical in nature and whether cumulative trauma resulted in PTSD.

**Methods:** We performed a 1:3 case-control study. The authors compared CVD patients diagnosed with PTSD (n=37) to those with adjustment disorder (n=111) in terms of trauma/stressor types and medical and demographic characteristics.

**Results:** Half (51%) of CVD patients suffering from PTSD had endured a medical trauma, 35% an external (non-medical) trauma, and 14% both. There were no significant differences with CVD patients diagnosed with adjustment disorder, 40% of them having experienced a medical stressor, 40% an external (non-medical) stressor and 20% both. Cumulative trauma was seen in only 19% of CVD patients suffering from PTSD. Traditional risk factors (female sex, younger age) were not prominent in CVD patients with PTSD as compared to those with adjustment disorder. Cases were, however, significantly more likely to have psychiatric antecedents and recent surgical interventions.

**Conclusions:** By uncovering characteristics of PTSD patients/trauma in CVD patients, this work will serve future research and clinical initiatives to better screen at-risk patients or at-risk medical situations.

---

While interpersonal and war trauma have been extensively studied as types of trauma associated with the development of posttraumatic stress disorder (PTSD) ^1-13^, medical conditions have only just recently begun garnering attention as trauma leading to PTSD ^14^. The concept of medical trauma was officially introduced in the fourth edition of the Diagnostic and Statistical Manual of Mental Disorders (DSM-IV). Interpretations of this definition varied widely within clinical and research fields, with some considering diagnosis of a severe, though not immediately life-threatening, illness, such as cancer, as possible trauma/stressor ^15^, while others only accepted conditions associated with an actual and immediate vital threat (such as myocardial infarction, stroke, or intensive care unit stay ^16-18^). This ambiguity was clarified in 2015, with DSM-5 stating that the medical condition need be sudden, unexpected, and potentially lethal in order to be considered as a possible PTSD trauma ^19^. The literature on PTSD and medical traumas remains sparse, but some studies report PTSD rates of up to 20% after significant medical trauma ^20,21^. Surprisingly, only a minority of these studies address the possibility that patients may have undergone both medical (e.g., massive myocardial infarction, defibrillator shock) and non-medical traumas (e.g., war, physical threat, or assault). Since experiencing cumulative traumas is a known PTSD risk factor ^22^, this gap substantially limits the interpretation and clinical applicability of the current literature on medical PTSD.

Within medical populations, there is long-standing recognition that cardiovascular diseases (CVD) are associated with PTSD ^23,24^. Research into these particular concurrently- associated comorbidities is complex as they share multiple risk factors for their development, such as an elevated heart rate, tobacco use, dyslipidemia, and obesity ^25^, and have a bidirectional relationship ^26,27^. It is still debated whether or not PTSD is an independent risk factor for the development of CVD ^28,29^, with PTSD oftentimes present concurrently in patients with CVD in large part due to the high prevalence of the latter disease. Specifically, the prevalence of heart diseases reached 10.6% in USA in recent years ^30^. As PTSD is associated with poorer cardiac prognosis ^31,32^, the co-occurrence of PTSD and CVD is not to be neglected. As a matter of fact, patients developing PTSD after an acute coronary syndrome double their risk of recurrent cardiac events and mortality compared to those without PTSD ^33^. Furthermore, patients suffering from CVD are at risk of being exposed to events that satisfy DSM-5 definitions of PTSD trauma/stressors such as incident or recurrent myocardial infarctions, malignant arrhythmias, and/or defibrillator shocks, thus concretizing the bidirectional nature and self-perpetuating cycle of PTSD and CVD mentioned above.

Whether a patient suffering from both PTSD and CVD was exposed to single or multiple medical traumas and/or to prior non-medical trauma(s) remains vastly unknown. To the best of our knowledge, the description of trauma types has never been studied in a cardiac population with comorbid PTSD. This is crucial to better prevent, screen, and treat PTSD within this significant medical population, thereby reducing this burdensome association.

A case-control study was thus performed to examine types of trauma associated with PTSD in a cardiac population. The hypothesis is that in CVD patients with PTSD: 1) medical trauma would be twice as frequent as non-medical trauma, and that 2) most patients will have been exposed to cumulative trauma (two or more).

## Methods

The Montreal Heart Institute (MHI) internal research and ethics committees approved this study.

We designed a case-control study to examine the nature of trauma associated with PTSD in CVD patients. A 1:3 ratio of CVD patients with diagnosed PTSD versus CVD patients with diagnosed adjustment disorder was selected given that such a ratio has been established to be efficient for optimisation of statistical power ^34^. Data was obtained by reviewing the medical charts of patients seen in the Cardio-Psychiatry Clinic of the MHI between January 1, 2013 and September 30, 2019. The MHI is a quaternary centre specialized in cardiovascular medicine and surgery based in Montreal, Quebec and affiliated with the Université de Montréal. The mental disorder diagnosis was obtained during a routine clinical psychiatric evaluation.

All case subjects were older than 18 years of age with a confirmed CVD and a full or partial diagnosis of PTSD. We considered PTSD full if the patient met all the DSM-IV or DSM-5 criteria and partial if the patient met some, but not all, of the criteria. Subjects with potential CVD under investigation, but not yet confirmed, and subjects whose medical charts were illegible were excluded. The control group consisted of subjects referred to the same outpatient clinic during the same period with a confirmed CVD and a diagnosis of adjustment disorder. The same exclusion criteria were applied to the control group.

We matched cases and controls (using MedCalc statistical software version 19.1.7) by the date of their clinic visit (± 120 days). This choice was made in order to limit the variability between psychiatrists, given that seven different psychiatrists worked at the clinic during the six-year study period.

Two trained staff members (a MSc candidate, MJM, and a PhD candidate, SC) performed data collection which consisted of obtaining demographic data, nature and amount of traumas/stressors, cardiac diseases, psychiatric comorbidity and treatment via thorough review of the patient’s medical records. The primary point of comparison, bearing in mind the study’s primary outcome, was the nature of the trauma/stressor associated with the psychiatric disorder (i.e., PTSD or adjustment disorder). Cases and controls were equally compared in terms of psychiatric antecedents, principal CVD, and previous hospitalizations or surgery and associated complications. Author KD, a cardiologist at the MHI, was consulted when a subject had multiple CVD and there was a resulting doubt about which should be considered the principal CVD. It was determined that the principal CVD in such cases would be the most clinically severe of the two (or more) CVD diagnoses. Three categories of CVD were established for the sake of the study: coronary artery disease, heart failure and/or valvular heart disease, and arrhythmia. The principal cardiac diagnosis should not be confounded with the medical trauma/stressor, which is defined as the event (e.g., myocardial infarction) associated with PTSD or adjustment disorder symptoms, regardless of its clinical importance

The authors chose to contrast the terms “medical” and “non-medical” to categorize the nature of trauma/stressors in CVD patients. Medical trauma/stressor was defined as an intracorporeal condition or disease, such as a myocardial infarction, arrhythmia, or reanimated cardiac arrest. Non-medical trauma/stressor was defined as originating from outside the body, such as environmental or social events (e.g., abuse, witnessing death of others). The term cumulative trauma/stressor was used if two or more traumas/stressors were associated with the development of PTSD or adjustment disorder. Subjects may have had two or more medical or non-medical traumas/stressors, or even potentially mixed (i.e., both medical and non-medical) traumas/stressors. Traumas needed to meet the DSM-5 definition (i.e., sudden, unexpected, and potentially lethal). These criteria were not required for an event to be classified as a stressor.

Socio-demographic characteristics, psychiatric and cardiovascular health comorbidities, and the nature of the event or trauma for cases and controls are presented. Continuous variables are presented as mean and standard deviation (SD) and categorical variables as count and percentages. Association between socio-demographic characteristics, comorbidities, the nature of the event, and the psychiatric disorder (i.e., PTSD or adjustment disorder) were tested for using logistic regression analysis. For categorical variables with a category for which one group had a count of 0, categories were either combined when appropriate or the Fisher’s test was employed. For all tests, a *p*-value <0.05 was considered significant. Statistical analyses were performed using SAS version 9.4 (SAS Institute Inc., Cary, North Carolina).

## Results

Of the 1,244 medical files reviewed, 20 patients were diagnosed with partial PTSD and 17 with full PTSD. By comparison, 257 patients were diagnosed with adjustment disorder prior to matching. To account for the desired 1:3 ratio, 37 cases were matched with 111 controls and further studied.

Demographic, medical, and surgical characteristics of cases and controls are shown in Table 1. There were no differences between the two groups in terms of age, sex, place of birth, occupational status, living arrangements (i.e., alone or with others), and civil status. However, patients with PTSD were more on medical leave compared to those with adjustment disorder (12 [38%] vs. 20 [20%], p=0.03). Patients with PTSD had also more psychiatric comorbidities (10 [27%] vs. 14 [13%], p=0.04) and were more likely to have a family history of psychiatric illness (13 [35%] vs. 14 [13%], p=0.003) compared to patients with adjustment disorder. The frequencies of the principal cardiac diagnoses (i.e., coronary heart disease, arrhythmia, and heart failure/valvular heart disease) as well as the proportions of patients with two or more cardiac diagnoses, pacemakers/defibrillators, hospitalization in the previous year, and medical/surgical complications were similar between both groups. However, the proportion of patients with surgery in the previous year was significantly higher in PTSD patients (11 [30%] vs. 13 [12%], p=0.01).

**Table 1.**
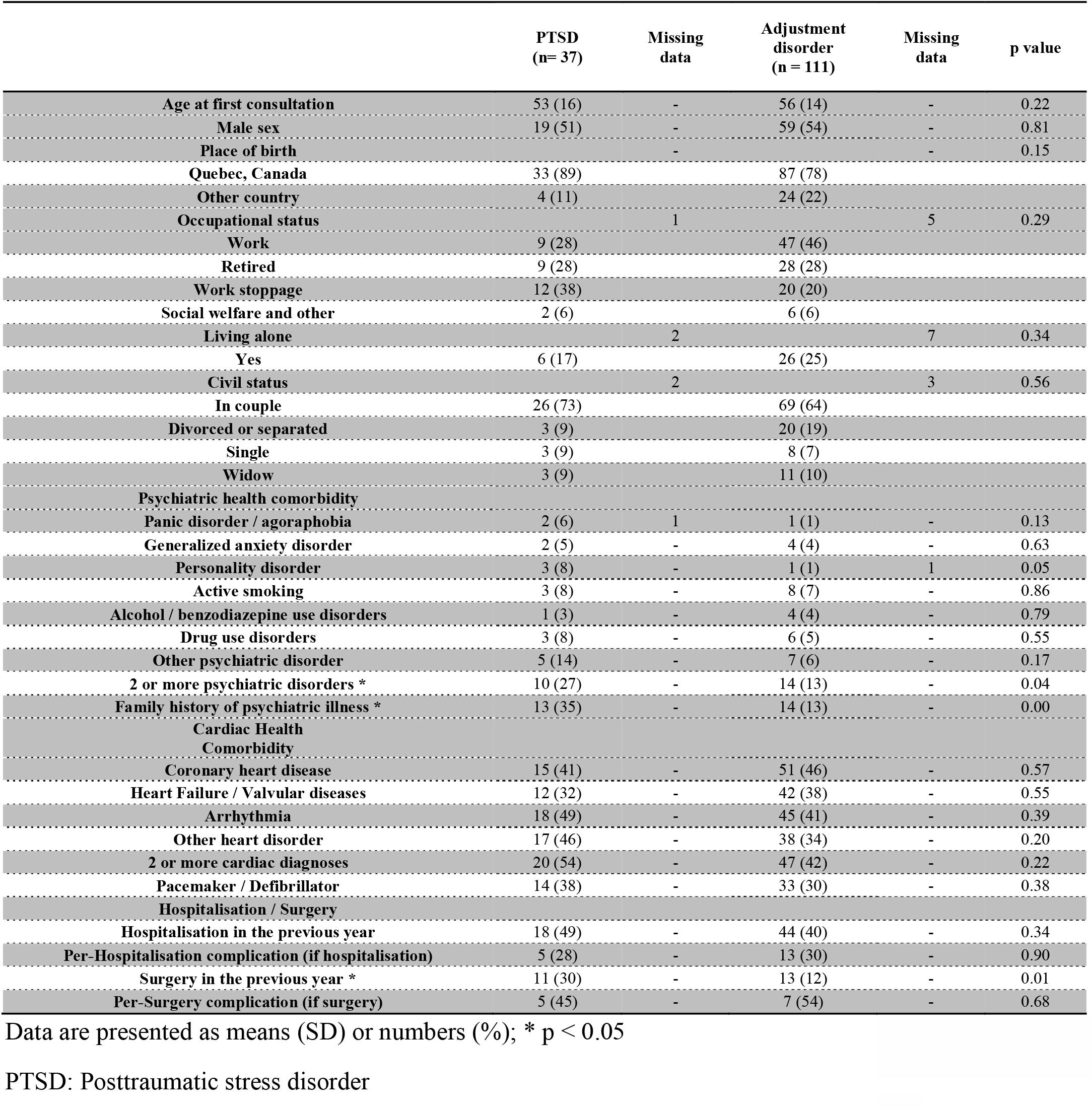
Socio demographic characteristics and psychiatric / cardiovascular health comorbidity of the study cases and controls

Table 2 describes the nature of trauma associated with PTSD development. Nineteen patients (51%) experienced a medical trauma, thirteen patients (35%) a non-medical trauma, and five (14%) both medical and non-medical trauma (1.3:1 medical to non-medical ratio). There was no significant difference (p=0.43) with the control group, with 40% having suffered a medical stressor, 40% a non-medical stressor, and 20% both types of stressors (1:1 medical to non-medical ratio). Only seven PTSD patients (19%) had cumulative traumas.

**Table 2.** Nature of the event associated with the psychiatric disorder and referral data in cardiopsychiatry clinic of study participants

|  | PTSD<br>(n = 37) | Missing<br>data | Adjustment<br>disorder<br>(n = 111) | Missing<br>data | p value | Adjustment<br>disorder exposed<br>to trauma<br>(n = 38) | Missing<br>data | p value |
| --- | --- | --- | --- | --- | --- | --- | --- | --- |
| **Event types** |  | - |  | - | 0.43 |  | - | 0.04 |
| **Medical** | 19 (51) |  | 44 (40) |  |  | 27 (71) |  |  |
| **External** | 13 (35) |  | 45 (40) |  |  | 11 (29) |  |  |
| **Medical and**<br>**External** | 5 (14) |  | 22 (20) |  |  | 0 (0) |  |  |
| **2 or more events** | 7 (19) | - | 37 (34) | 3 | 0.08 | 4 (11) | - | 0.72 |
| **Medical event**<br>**type, if applicable** |  | - |  | - |  |  | - |  |
| **Myocardial**<br>**infarction** | 7 (29) |  | 8 (12) |  |  | 8 (30) |  |  |
| **Arrhythmia** | 6 (25) |  | 4 (6) |  |  | 4 (15) |  |  |
| **Inappropriate**<br>**defibrillator**<br>**shock** | 1 (4) |  | 4 (6) |  |  | 3 (11) |  |  |
| **Other** | 10 (42) |  | 50 (76) |  |  | 12 (44) |  |  |
| **External event**<br>**type, if applicable** |  | - |  | - |  |  | - |  |
| **Interpersonal** | 13 (72) |  | 66 (98) |  |  | 11 (100) |  |  |
| **War and**<br>**Witnessing** | 5 (28) |  | 1 (2) |  |  | 0 (0) |  |  |
| **Location of the**<br>**event** |  | 1 |  | 6 | 0.95 |  | 2 | 0.18 |
| **Outside of the**<br>**hospital** | 28 (76) |  | 80 (76) |  |  | 22 (61) |  |  |
| **Inside the walls of**<br>**the hospital** | 9 (24) |  | 25 (24) |  |  | 14 (39) |  |  |

Data are presented as means (SD) or numbers (%)
PTSD: Posttraumatic stress disorder

Medical traumas suffered by patients diagnosed with PTSD were mainly myocardial infarctions (29%), malignant arrhythmia (25%), and defibrillator shocks (4%). Other types of medical traumas (regrouping the remaining 42% of PTSD patients) were multi-complicated heart surgery, intensive care unit stays, and cerebrovascular accidents. With regards to non-medical traumas in patients with PTSD, 72% were interpersonal (aggression, rape, etc.), 22% had witnessed a traumatic situation, and 6% had participated in a war.

## Discussion

Contrary to the hypothesis that trauma would mostly be medical in nature in patients diagnosed with PTSD in a quaternary cardiac hospital, a ratio close to 1:1 in terms of medical to non-medical trauma types was observed in this study. This suggests that, even in a highly medical population, health issues are but one of many possible human traumas/stressors, thereby reinforcing the importance of holistic, bio-psycho-social patient care rather than a simple focus on physical ailments and medical stressors. This study lends credence to the fact that CVD may have a low traumatic potential ^35^. Although CVD is associated with its share of social stigma ^36^, it may be less stigmatized than non-medical trauma (e.g., rape ^37^). Consequently, patients with CVD tend to more naturally utilize their social support network, which is known to have a protective effect against the development of PTSD ^38^, thereby potentially contributing to the less psychologically traumatic nature of CVD disease.

PTSD and adjustment disorder patients were similar in terms of principal and number of cardiac diseases, presence of a pacemaker/defibrillator, prior hospitalization, and past medical complications. This is consistent with previous works and existing literature on medical PTSD, which has failed to find an association between PTSD risk and the severity of pathology or CVD type ^35,39-41^.

However, this study’s data did reveal one medical difference between PTSD and adjustment disorder: surgery in the previous year was more frequent in PTSD patients than in adjustment disorder patients. It is known that the traumatic nature of a medical event pertains more closely to the patient’s (“subjective”) perception of a death threat than to the medical (“objective”) severity of the illness ^42^. Cardiovascular surgery, whose major nature, risks of complications, and subsequent ICU stay may certainly appear a death threat to patients, harmonizes the aforementioned principles and very likely explain the difference noted between PTSD and adjustment disorder patients having undergone surgery in the past year. Surprisingly, cumulative trauma was only seen in 19% of CVD patients suffering from PTSD.

Traditionally recognized risk factors for PTSD development, such as female sex and younger age ^19^, were not evident in this study’s CVD population. This may be explained by the fact that risk factors for CVD are opposite to those of PTSD in terms of sex and age ^43-46^. Indeed, CVD patients are often older males whereas PTSD patients tend to be younger females. This study’s data suggest that personal and family history of psychiatric diseases, as well as medical leave from work, would be more informative risk factors for PTSD development in the CVD population.

One of the most important characteristics of a case-control study is the choice of controls, ^47^ with controls being theoretically at risk of becoming a case. We selected patients with adjustment disorder as our control group given that, largely, they experienced similar medical/non-medical traumas to PTSD patients, though did not themselves go on to develop PTSD. DSM-5 classifies adjustment disorder, which arises from difficulty adjusting or coping following a stressor, within the same section as PTSD, namely Trauma- and Stressor-Related Disorders. Both cases and controls were possibly subjected to similar trauma of both medical and non-medical types.

Although the concept of cumulative trauma is widely recognized with the psychiatric community ^48,49^, it is still possible that psychiatrists did not systematically ask about pre-existing traumatic experiences. This is a limit of this retrospective study of medical files, but does represent real-life psychiatric interviews, free of voluntary bias associated with prospective cohort studies. It could also be argued that including partial PTSD weakens the internal validity, but because patients can display different symptomatic presentations along the PTSD spectrum, their inclusion is in line with the now well-recognized dimensional classification of psychiatric disorders ^50-54^.

Regarding non-medical validity, it is possible that patients suffering from PTSD but not referred to the Cardio-Psychiatric Clinic bear different characteristics and traumatic stories than the ones included in the present study. Since one of the main symptoms of PTSD is avoidance, notably of the place where the trauma occurred, it is possible that this study is blinded to the most severe cases of PTSD and to those avoiding returning to the site of their trauma. A sub-analysis with adjustment disorder patients for whom stressors fit the trauma definition was performed. In those patients, the distribution of event locations was like that of PTSD patients, thus invalidating the previous hypothesis. Finally, generalization to other clinical settings should be done with caution, as the data was derived from a quaternary cardiac hospital.

Cardiology professionals ought to consider evenly both medical and non-medical events as sources of possible psychological distress in their CVD patients. They should also be aware that traditional PTSD risk factors (i.e., female sex and younger age) do not apply to CVD patients, who tend to be older and male. It would be beneficial to rather pay special attention to the mental status of patients on medical leave, with a personal or family history of psychiatric diseases, and/or those having undergone surgery within the last year. Those sharing sustained psychological distress or PTSD symptoms should be referred to mental health community services or specialists.

In research, prospective studies may be of interest for identifying modifiable risk and protective factors for PTSD development in high-risk CVD population (i.e., upcoming surgery, being on medical leave, having personal or family history of psychiatric diseases). The goal is early detection of patients at risk, thus enabling prevention via the offering of resources/treatments. This is of primal importance given that the PTSD-CVD combination is a very burdensome comorbidity to carry both in terms of psychological suffering and poor evolution of CVD ^55^.

## Data Availability

All data will be made available upon request.

## Acknowledgments

The authors thank Montreal Heart Institute Foundation for the funding granted to Dre. Judith Brouillette.

## References

1. Creamer M, Burgess P, McFarlane AC. Post-traumatic stress disorder: findings from the Australian National Survey of Mental Health and Well-being. Psychol Med 2001;31:1237–1247.

2. Donoho CJ, Bonanno GA, Porter B, Kearney L, Powell TM. A Decade of War: Prospective Trajectories of Posttraumatic Stress Disorder Symptoms Among Deployed US Military Personnel and the Influence of Combat Exposure. Am J Epidemiol 2017;186:1310–1318.

3. Hoge CW, Terhakopian A, Castro CA, Messer SC, Engel CC. Association of posttraumatic stress disorder with somatic symptoms, health care visits, and absenteeism among Iraq war veterans. Am J Psychiatry 2007;164:150–153.

4. Lawson NR. Posttraumatic stress disorder in combat veterans. JAAPA 2014;27:18–22.

5. Muller J, Ganeshamoorthy S, Myers J. Risk factors associated with posttraumatic stress disorder in US veterans: A cohort study. PloS one 2017;12:e0181647.

6. Phillips EL. War neurosis; a preliminary study of clinical and control groups. J Clin Psychol 1947;3:148–154.

7. Phillips EL. War neurosis; a second study of clinical and control goups. J Clin Psychol 1947;3:155–164.

8. Richardson LK, Frueh BC, Acierno R. Prevalence estimates of combat-related post-traumatic stress disorder: critical review. Aust N Z J Psychiatry 2010;44:4–19.

9. Selinski H. Traumatic neurosis of war. J Indiana State Med Assoc 1946;39:170.

10. Spiegel JP, Grinker RR. War neuroses. Prog Neurol Psychiatry 1946;1:579–596.

11. Stern RL. Diary of a war neurosis. J Nerv Ment Dis 1947;106:583–586.

12. Thomas JL, Wilk JE, Riviere LA, McGurk D, Castro CA, Hoge CW. Prevalence of mental health problems and functional impairment among active component and National Guard soldiers 3 and 12 months following combat in Iraq. Arch Gen Psychiatry 2010;67:614–623.

13. Walsh K, Koenen KC, Aiello AE, Uddin M, Galea S. Prevalence of sexual violence and posttraumatic stress disorder in an urban African-American population. J Immigr Minor Health 2014;16:1307–1310.

14. Mundy E, Baum A. Medical disorders as a cause of psychological trauma and posttraumatic stress disorder. Curr Opin Psychiatr 2004;17:123–127.

15. Swartzman S, Booth JN, Munro A, Sani F. Posttraumatic stress disorder after cancer diagnosis in adults: A meta-analysis. Depression and anxiety 2017;34:327–339.

16. Lima BB, Hammadah M, Wilmot K, Pearce BD, Shah A, Levantsevych O, Kaseer B, Obideen M, Gafeer MM, Kim JH, Sullivan S, Lewis TT, Weng L, Elon L, Li L, Bremner JD, Raggi P, Quyyumi A, Vaccarino V. Posttraumatic stress disorder is associated with enhanced interleukin-6 response to mental stress in subjects with a recent myocardial infarction. Brain Behav Immun 2019;75:26–33.

17. Rutovic S, Kadojic D, Dikanovic M, Solic K, Malojcic B. Prevalence and correlates of post-traumatic stress disorder after ischaemic stroke. Acta Neurol Belg 2019.

18. Wu KK, Cho VW, Chow FL, Tsang AP, Tse DM. Posttraumatic Stress after Treatment in an Intensive Care Unit. East Asian arch 2018;28:39–44.

19. American Psychiatric A, Benyamina A, Guilabert C, Guelfi J-D, Crocq M-A, Boyer P, Pull C-B, Pull M-C, Abbar M, Arbabzadeh-Bouchez S. DSM-5 - Manuel Diagnostique et Statistique des Troubles Mentaux. Philadelphia, PA, FRANCE: Elsevier - Health Sciences Division, 2015.

20. Ginzburg K, Ein-Dor T. Posttraumatic stress syndromes and health-related quality of life following myocardial infarction: 8-year follow-up. Gen Hosp Psychiatry 2011;33:565–571.

21. Ingles J, Sarina T, Kasparian N, Semsarian C. Psychological wellbeing and posttraumatic stress associated with implantable cardioverter defibrillator therapy in young adults with genetic heart disease. Int J Cardiol 2013;168:3779–3784.

22. Ogle CM, Rubin DC, Siegler IC. Cumulative exposure to traumatic events in older adults. Aging Ment Health 2014;18:316–325.

23. Edmondson D, Cohen BE. Posttraumatic stress disorder and cardiovascular disease. Prog Cardiovasc Dis 2013;55:548–556.

24. Deng LX, Khan AM, Drajpuch D, Fuller S, Ludmir J, Mascio CE, Partington SL, Qadeer A, Tobin L, Kovacs AH, Kim YY. Prevalence and Correlates of Post-traumatic Stress Disorder in Adults With Congenital Heart Disease. Am J Cardiol 2016;117:853–857.

25. Dyball D, Evans S, Boos CJ, Stevelink SAM, Fear NT. The association between PTSD and cardiovascular disease and its risk factors in male veterans of the Iraq/Afghanistan conflicts: a systematic review. Int Rev Psychiatry 2019;31:34–48.

26. Carmassi C, Cordone A, Pedrinelli V, Dell’Osso L. PTSD and Cardiovascular Disease. In: Govoni S, Politi P, Vanoli E, eds. Brain and Heart Dynamics. Cham: Springer International Publishing, 2020:1–23.

27. De Hert M, Detraux J, Vancampfort D. The intriguing relationship between coronary heart disease and mental disorders. Dialogues Clin Neurosci 2018;20:31–40.

28. Edmondson D, Kronish IM, Shaffer JA, Falzon L, Burg MM. Posttraumatic stress disorder and risk for coronary heart disease: a meta-analytic review. Am Heart J 2013;166:806–814.

29. Scherrer JF, Salas J, Cohen BE, Schnurr PP, Schneider FD, Chard KM, Tuerk P, Friedman MJ, Norman SB, van den Berk-Clark C, Lustman PJ. Comorbid Conditions Explain the Association Between Posttraumatic Stress Disorder and Incident Cardiovascular Disease. J Am Heart Assoc 2019;8:e011133.

30. Virani SS, Alonso A, Benjamin EJ, Bittencourt MS, Callaway CW, Carson AP, Chamberlain AM, Chang AR, Cheng S, Delling FN, Djousse L, Elkind MSV, Ferguson JF, Fornage M, Khan SS, Kissela BM, Knutson KL, Kwan TW, Lackland DT, Lewis TT, Lichtman JH, Longenecker CT, Loop MS, Lutsey PL, Martin SS, Matsushita K, Moran AE, Mussolino ME, Perak AM, Rosamond WD, Roth GA, Sampson UKA, Satou GM, Schroeder EB, Shah SH, Shay CM, Spartano NL, Stokes A, Tirschwell DL, VanWagner LB, Tsao CW, American Heart Association Council on E, Prevention Statistics C, Stroke Statistics S. Heart Disease and Stroke Statistics-2020 Update: A Report From the American Heart Association. Circulation 2020;141:e139–e596.

31. Burg MM, Soufer R. Post-traumatic Stress Disorder and Cardiovascular Disease. Curr Cardiol Rep 2016;18:94.

32. Cavalcanti-Ribeiro P, Andrade-Nascimento M, Morais-de-Jesus M, de Medeiros GM, Daltro-Oliveira R, Conceicao JO, Rocha MF, Miranda-Scippa A, Koenen KC, Quarantini LC. Post-traumatic stress disorder as a comorbidity: impact on disease outcomes. Expert Rev Neurother 2012;12:1023–1037.

33. Edmondson D, Richardson S, Falzon L, Davidson KW, Mills MA, Neria Y. Posttraumatic stress disorder prevalence and risk of recurrence in acute coronary syndrome patients: a meta-analytic review. PloS one 2012;7:e38915.

34. Hennessy S, Bilker WB, Berlin JA, Strom BL. Factors influencing the optimal control-to-case ratio in matched case-control studies. Am J Epidemiol 1999;149:195–197.

35. Roberge MA, Dupuis G, Marchand A. Post-traumatic stress disorder following myocardial infarction: prevalence and risk factors. The Canadian journal of cardiology 2010;26:e170–175.

36. Panza GA, Puhl RM, Taylor BA, Zaleski AL, Livingston J, Pescatello LS. Links between discrimination and cardiovascular health among socially stigmatized groups: A systematic review. PloS one 2019;14:e0217623.

37. Deitz MF, Williams SL, Rife SC, Cantrell P. Examining cultural, social, and self-related aspects of stigma in relation to sexual assault and trauma symptoms. Violence Against Women 2015;21:598–615.

38. Simon N, Roberts NP, Lewis CE, van Gelderen MJ, Bisson JI. Associations between perceived social support, posttraumatic stress disorder (PTSD) and complex PTSD (CPTSD): implications for treatment. Eur J Psychotraumatol 2019;10:1573129.

39. Kapa S, Rotondi-Trevisan D, Mariano Z, Aves T, Irvine J, Dorian P, Hayes DL. Psychopathology in patients with ICDs over time: results of a prospective study. Pacing and clinical electrophysiology : PACE 2010;33:198–208.

40. Habibovic M, Denollet J, Pedersen SS, on behalf of the Wi. Posttraumatic stress and anxiety in patients with an implantable cardioverter defibrillator: Trajectories and vulnerability factors. Pacing and clinical electrophysiology : PACE 2017;40:817–823.

41. Long V, Guertin MC, Dyrda K, Benrimoh D, Brouillette J. Descriptive Study of Anxiety and Posttraumatic Stress Disorders in Cardiovascular Disease Patients: From Referral to Cardiopsychiatric Diagnoses. Psychother Psychosom 2018;87:370–371.

42. Sareen J. Posttraumatic stress disorder in adults: impact, comorbidity, risk factors, and treatment. Can J Psychiatry 2014;59:460–467.

43. D’Agostino RB, Sr. Vasan RS, Pencina MJ, Wolf PA, Cobain M, Massaro JM, Kannel WB. General cardiovascular risk profile for use in primary care: the Framingham Heart Study. Circulation 2008;117:743–753.

44. Kappert K, Bohm M, Schmieder R, Schumacher H, Teo K, Yusuf S, Sleight P, Unger T, Investigators OT. Impact of sex on cardiovascular outcome in patients at high cardiovascular risk: analysis of the Telmisartan Randomized Assessment Study in ACE-Intolerant Subjects With Cardiovascular Disease (TRANSCEND) and the Ongoing Telmisartan Alone and in Combination With Ramipril Global End Point Trial (ONTARGET). Circulation 2012;126:934–941.

45. Savji N, Rockman CB, Skolnick AH, Guo Y, Adelman MA, Riles T, Berger JS. Association between advanced age and vascular disease in different arterial territories: a population database of over 3.6 million subjects. J Am Coll Cardiol 2013;61:1736–1743.

46. Tunstall-Pedoe H, Kuulasmaa K, Mahonen M, Tolonen H, Ruokokoski E, Amouyel P. Contribution of trends in survival and coronary-event rates to changes in coronary heart disease mortality: 10-year results from 37 WHO MONICA project populations. Monitoring trends and determinants in cardiovascular disease. Lancet 1999;353:1547–1557.

47. Lewallen S, Courtright P. Epidemiology in practice: case-control studies. Community Eye Health 1998;11:57–58.

48. Briere J, Agee E, Dietrich A. Cumulative trauma and current posttraumatic stress disorder status in general population and inmate samples. Psychol Trauma 2016;8:439–446.

49. Cloitre M, Stolbach BC, Herman JL, van der Kolk B, Pynoos R, Wang J, Petkova E. A developmental approach to complex PTSD: childhood and adult cumulative trauma as predictors of symptom complexity. J Trauma Stress 2009;22:399–408.

50. First MB. Clinical utility: a prerequisite for the adoption of a dimensional approach in DSM. J Abnorm Psychol 2005;114:560–564.

51. Widiger TA, Samuel DB. Diagnostic categories or dimensions? A question for the Diagnostic And Statistical Manual Of Mental Disorders--fifth edition. J Abnorm Psychol 2005;114:494–504.

52. Helzer JE, Kraemer HC, Krueger RF. The feasibility and need for dimensional psychiatric diagnoses. Psychol Med 2006;36:1671–1680.

53. Kraemer HC. DSM categories and dimensions in clinical and research contexts. Int J Methods Psychiatr Res 2007;16 Suppl 1:S8–S15.

54. Regier DA. Dimensional approaches to psychiatric classification: refining the research agenda for DSM-V: an introduction. Int J Methods Psychiatr Res 2007;16 Suppl 1:S1–5.

55. Coughlin SS. Post-traumatic Stress Disorder and Cardiovascular Disease. Open Cardiovasc Med J 2011;5:164–170.

